# Quantifying ^18^F-Fluorodeoxyglucose Uptake in Perianal Fistulas on PET/CT: A Retrospective Analysis

**DOI:** 10.1101/2023.11.07.23298231

**Authors:** Katherine Huang, Favour Garuba, Aravinda Ganapathy, Grace Bishop, Hanjing Zhang, Adriene Lovato, Malak Itani, Satish E. Viswanath, Tyler J. Fraum, Parakkal Deepak, David H. Ballard

**Affiliations:** Division of Gastroenterology, Washington University School of Medicine in St. Louis, St. Louis, MO, USA; Washington University School of Medicine in St. Louis, St. Louis, MO, USA; Mallinckrodt Institute of Radiology, Washington University School of Medicine in St. Louis, St. Louis, MO, USA; Department of Biomedical Engineering, School of Engineering, Case Western Reserve University, Cleveland, Ohio, USA

## Abstract

**Rationale and Objectives:** The use of ^18^F-fluorodeoxyglucose positron emission tomography-computed tomography (FDG-PET/CT) in assessing inflammatory diseases has shown significant promise. Uptake patterns in perianal fistulas, which may be an incidental finding on PET/CT, have not been purposefully studied. Our aim was to compare FDG uptake of perianal fistulas to that of the liver and anal canal in patients who underwent PET/CT for hematologic/oncologic diagnosis or staging.

**Materials and Methods:** We retrospectively identified patients who underwent FDG-PET/CT imaging between January 2011 and May 2023, where the report described a perianal fistula or abscess. PET/CTs of patients included in the study were retrospectively analyzed to record the maximum standardized uptake value (SUV_max_) of the fistula, abscess, anal canal, rectum, and liver. Fistula-to-liver and Fistula-to-anus SUV_max_ ratios were calculated. We statistically compared FDG activity among the fistula, liver, and anal canal. We also assessed FDG activity in patients with vs. without anorectal cancer, as well as across different St. James fistula grades.

**Results:** The study included 24 patients with identifiable fistulas. Fistula SUV_max_ (mean=10.8±5.28) was significantly higher than both the liver (mean=3.09±0.584, p<0.0001) and the anal canal (mean=5.98± 2.63, p=0.0005). Abscess fistula SUV_max_ was 15.8 ± 4.91. St. James grade 1 fistulas had significantly lower SUV_max_ compared to grades 2 and 4 (p= 0.0224 and p=0.0295 respectively). No significant differences existed in SUV_max_ ratios between anorectal and non-anorectal cancer groups.

**Conclusion:** Perianal fistulas have increased FDG avidity with fistula SUV_max_ values that are significantly higher than the anal canal.

## Introduction

The prevalence of perianal fistulas is estimated to be 1.69 per 10,000 with an annual incidence of 1.15 per 10,000. Cryptoglandular infection and Crohn’s disease account for 50.8% and 44.6% of prevalent cases, respectively. Other etiologies, such as infection, trauma, and cancer, account for the remaining 5% of fistula cases [1].

Immunosuppressed patients undergoing chemotherapy are prone to develop perianal fistulas and abscesses due to compromised immune responses and altered inflammatory processes. These perianal fistulas and abscesses impact patient outcomes and quality of life, necessitating accurate detection and monitoring.

Magnetic Resonance Imaging (MRI) is typically utilized for the assessment of perianal fistulas due to its excellent soft tissue resolution which allows for high sensitivity and specificity in detecting fistulous tracts [2, 3]. Conventional CT can be utilized in the detection of anorectal abscesses but performs inferiorly to MRI in the assessment of fistulas due to its limited resolution which leads to suboptimal visualization and delayed diagnosis [4, 5]. Utilizing ^18^F-fluorodeoxyglucose positron emission tomography-computed tomography (FDG-PET/CT) may provide an avenue to overcome these challenges by providing additional functional information about the anorectal region, which could enhance the sensitivity and specificity of detecting and characterizing these anorectal pathologies with CT. Moreover, perianal fistulas may be encountered as an incidental finding on PET/CT performed for oncologic staging and diagnosis.

Over the past two decades, FDG-PET/CT has emerged as a valuable tool for the assessment of infectious and inflammatory disorders and their response to treatment. The application of FDG-PET/CT to various infectious and inflammatory disorders continues to increase with studies demonstrating its clinical utility in assessing chronic inflammatory bowel diseases such as luminal manifestations in Crohn’s disease [6, 7]. Studies using FDG-PET to analyze benign anorectal pathologies have been limited to hemorrhoids with no prior studies on FDG uptake in perianal fistulas and abscesses [8]. Additionally, recent changes (∼2021) in the policies of the Centers for Medicare and Medicaid Services (CMS) favor broader coverage for the use of FDG-PET for inflammation and infection as reflected in Society of Nuclear Medicine and Molecular Imaging (SNMMI) guidelines [9, 10].

Given the growing uses of FDG-PET/CT in infection and inflammation, we aimed to determine the clinical utility of FDG-PET/CT in imaging perianal fistulae and abscesses. The purpose of our study was to compare the pattern of FDG uptake in perianal fistulas to that of the liver and anal canal in patients who underwent PET/CT for hematologic and oncologic indications. A secondary objective was to assess for differences in fistula FDG uptake in patients with and without anorectal cancer and between different grades of fistulas based on St. James Grade.

## Materials and Methods

We conducted a single-center retrospective study approved by our Institutional Review Board. All study procedures were in accordance with the Health Insurance Portability and Accountability Act. The need for informed consent was waived due to the retrospective design (local IRB approval number 202204095). We retrospectively analyzed consecutive patients who underwent FDG-PET/CT imaging between January 2011 to May 2023 using a radiology information system (RIS) text search, which queried the clinical dictation for a description of a perianal fistula or abscess per details below, followed by image analysis of the FDG-PET/CT data.

Our RIS was queried using Montage (Montage 2023, Nuance, Burlington, MA, USA) for patients with an incidental perianal fistula or abscess. The search was limited to examinations coded and categorized as PET, grouped by individual patients, with the following structured search terminology: (“perianal fistula” | “in ano” | “anal fistula” | “rectal fistula” | “perianal tract” | “anal tract” | “perianal abscess” | “perirectal abscess” | “anorectal fistula” | “anorectal abscess”). Two study investigators (non-radiologists) assessed PET/CT reports of the initial subset of patients from the Montage search to exclude patients without a confirmed diagnosis of a perianal fistula or abscess, such as examinations that the report did not indicate a positive finding of perianal fistula or abscess (e.g., result returned because the report states “no evidence of residual perianal abscess as demonstrated on comparison CT”, or the history mentions one of the key search terms, but the findings or impression did not query this). Two study radiologists reviewed the PET/CT images of the remaining patients to exclude patients with fistulas that could not be well visualized and patients with anorectal cancer where the malignancy was indistinguishable from the perianal fistula. Tracer type and concurrently acquired imaging for fusion (i.e., PET/CT or PET/MR) were not search criteria; however, the cohort for potential inclusion only yielded results for FDG-PET/CT.

If patients had multiple PET/CTs that described a fistula within the study period, the earliest PET/CT that preceded a clinic visit or surgical procedure with provider notes that indicated the presence of a fistula was included for radiologist review. For patients without a clinic visit or surgical procedure, the first PET/CT that described the perianal fistula in the report was utilized. Patients were dichotomized as patients with versus without an anorectal malignancy (e.g., anal squamous cell carcinoma, rectal adenocarcinoma, or perianal Crohn’s disease with cancer in the fistulous tract) or perineal primary malignancy (e.g., vulvar cancer). Patients’ charts were systematically reviewed to document demographic information and relevant clinical records including relevant physical examination findings, concurrent imaging studies, and surgical procedures within 3 months of the reference FDG PET/CT scan date. A stepwise confirmation for perianal fistula was utilized (perianal fistula/abscess procedure > colorectal surgery clinical physical exam > other provider notes like a hospitalist note indicating signs or symptoms of a perianal fistula or abscess) to assess for a physician confirmatory diagnosis of a perianal fistula or abscess. Dedicated follow-up and fistula-related outcomes were not systematically assessed. No patients had administration of metformin prior to the PET.

Two study investigators, trained and supervised by a study radiologist, segmented the following: i) fistula or abscess, ii) the anal canal, iii) the rectum, and iv) a spherical region of interest on the right hemiliver (with a diameter of at least 3 cm). A study radiologist, an abdominal radiologist with 2 years post-fellowship experience, reviewed the patients identified for potential inclusion and the segmentation of the two study investigators. All imaging review was completed on a PACS interface (Sectra IDS7 version 25.1, Linköping, Sweden). The study radiologist reviewed (and adjusted, as necessary) segmentations of the regions of interest. The FDG fused state scale was modified to within 2 units of the identified SUV_max_ to ensure that the spherical volume-of-interest (VOI) was placed at the most FDG avid region of the structure of interest. A perianal fistula was defined as a linear FDG avid tract that extends from or involves the anal sphincter complex. Distinguishing a widely patent fistula from an abscess lacks a universally agreed-upon definition. In this study, we defined an abscess as a distinct, rounded area of attenuation separate from the surrounding subcutaneous or deep anatomical compartment fat or musculature, with a minimum diameter of 10 mm on the CT portion of the images. For the anus, the most FDG avid area not directly abutting the fistulous tract was utilized for the VOI SUV_max_ analysis. The rectum was defined as the bowel immediately above the puborectalis muscle (anorectal junction) and up to the peritoneal reflection (rectosigmoid junction). The study radiologist review focused on avoiding excreted FDG within the urinary bladder. In addition to validating and adjusting the VOI placement for perianal fistulas and abscesses, the study radiologist also classified the fistulas according to St. James and Parks fistula classification systems [11, 12]. After adjusting the segmentation and validating or excluding the patient, the final cohort for analysis was identified.

In our primary statistical analysis, we evaluated the ratio of the SUV_max_ of the fistulous tract to the SUV_max_ of the liver and anal canal. The SUV_max_ of the fistulous tract was categorized into five groups relative to that of the liver:

I. Minimal-Mild: SUV_max_ of the fistulous tract less than or equal to the SUV_max_ of the liver.
II. Moderate: SUV_max_ of the fistulous tract greater than the SUV_max_ of the liver but less than or equal to twice the SUV_max_ of the liver.
III. Moderate to marked: SUV_max_ of the fistulous tract greater than twice the SUV_max_ of the liver but less than or equal to thrice the SUV_max_ of the liver
IV. Marked: SUV_max_ of the fistulous tract greater than three times the SUV_max_ of the liver.

Patients were dichotomized into two subgroups based on cancer diagnosis. All patients with anorectal cancer were placed in one group while all patients without anorectal were placed in another group. SUV_max_ and SUV_max_ ratios were compared between both groups via statistical analysis. SUV_max_ of abscesses were recorded and analyzed separately from that of the fistulae.

The mean and standard deviation (SD) of SUV_max_ was calculated for all regions of interest including the fistula, abscess, liver, rectum, and anus. Statistical analyses explored relationships between the SUV_max_ of the perianal fistula and the liver and anal canal using paired t-tests. The mean statistical analyses were performed using GraphPad Prism version 10.0.0 for Windows, GraphPad Software, Boston, Massachusetts USA, www.graphpad.com, and Rstudio version 2023.06.1 [13]. Normality was assessed using the Shapiro-Wilk test. 1-way ANOVA with post-hoc Fisher’s Least Significant Difference test was used for significance testing in normally distributed variables, and Kruskal-Wallis rank sum test with post-hoc Dunn’s multiple comparison test was used for significance testing in non-normally distributed variables. A p-value of less than 0.05 was considered significant. For normal distributions, an unpaired t-test was used in cases of just 2 groups, and Mann-Whitney U test was used in cases of non-normal distribution.

## Results

Based on the structured PET dictation search, 63 patients were initially identified for this study. Following the dictation review, 34 patients were considered suitable for inclusion by two study investigators. Among these patients, 21 were non-anorectal cancer cases, and 4 were excluded due to the absence of fistula evidence during radiologist’s review of the PET/CT images. The remaining 13 had anorectal cancer and 6 were excluded due to the study radiologist’s inability to differentiate tumor from fistula. There were no patients with perineal cancer identified for inclusion in the study. Consequently, the final study cohort comprised of 24 eligible patients, including 17 without anorectal cancer and 7 with anorectal cancer. The study flow chart is summarized in **Figure 1**. Of note, 4 patients had established diagnosis of Crohn’s disease, all of whom were part of the anorectal cancer subgroup. Patient demographics, including age, sex, history of inflammatory bowel disease, clinical reference of perianal disease, and hematologic/oncologic diagnosis, are shown in **Table 1**.

**Table 1.** Patient demographics

|  | All Patients<br>(n=24) | Anorectal Group<br>(n=7) | Non-anorectal<br>Group (n=17) |
| --- | --- | --- | --- |
| Mean Age, years (range) | 56 (18-81) | 45 (34-63) | 60 (18-81) |
| Males, n (%) | 15 (62.5%) | 3 (42.9%) | 12 (70.6%) |
| Females, n (%) | 9 (37.5%) | 4 (57.1%) | 5 (29.4%) |
| History of inflammatory bowel disease, n | 4 | 4 | 0 |
| Clinical reference of perianal disease |  |  |  |
| Surgery, n (%) | 13 (54.1%) | 3 (42.9%) | 10 (58.8%) |
| Clinic visit, n (%) | 4 (16.7%) | 2 (28.6%) | 3 (17.6%) |
| Provider Notes, n (%) | 2 (8.3%) | 1 (14.3%) | 1 (5.9%) |
| None*, n (%) | 4 (20.8%) | 1 (14.3%) | 3 (17.6%) |
| Hematologic/Oncologic Diagnosis |  |  |  |
| Anorectal cancer |  |  |  |
| Anal, n (%) | 6 (25.0%) |  |  |
| Rectal, n (%) | 1 (4.2%) |  |  |
| Non-anorectal cancer |  |  |  |
| Lymphoma, n (%) | 7 (29.2%) |  |  |
| Lung, n (%) | 4 (16.7%) |  |  |
| Breast, n (%) | 2 (8.3%) |  |  |
| Plasmacytoma, n (%) | 1 (4.2%) |  |  |
| Esophageal, n (%) | 1 (4.2%) |  |  |
| Gastric, n (%) | 1 (4.2%) |  |  |
| Aplastic Anemia, n (%) | 1 (4.2%) |  |  |

\*No clinical reference of perianal disease within 3 months of PET accessible through our health care portal

**Figure 1.**
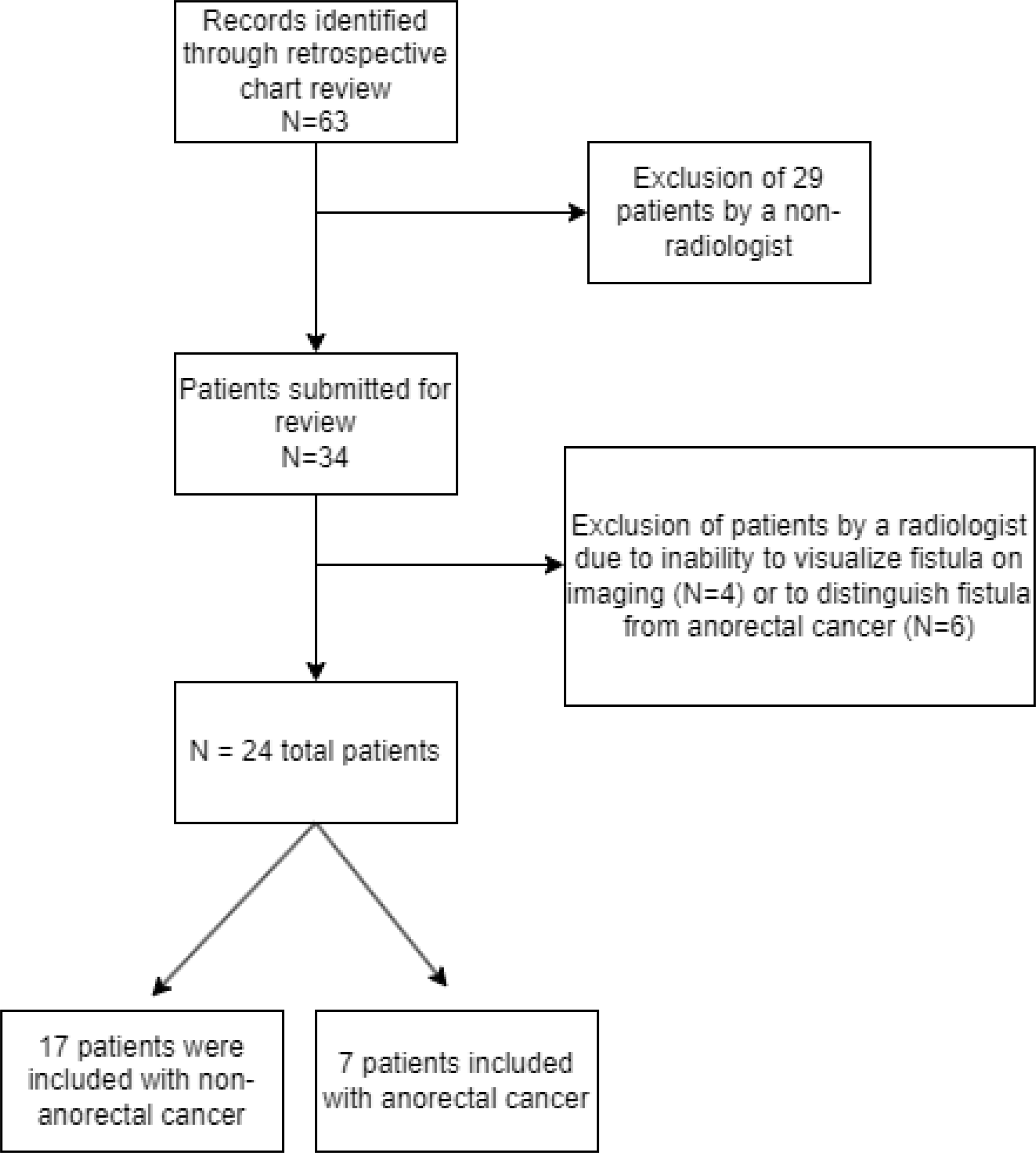
Study flow chart

Fistulas demonstrated higher average SUV_max_ (mean SUV_max_ =10.8±5.28) than the liver (mean SUV_max_ =3.09±0.584, p<0.0001) and the anus (mean SUV_max_ = 5.98± 2.63, p = 0.0005) (**Figure 2**).). A case example is denoted in **Figure 3**. For the 6 patients who concurrently had a perianal abscess, the average abscess SUV_max_ was 15.8±4.91.

**Figure 2.**
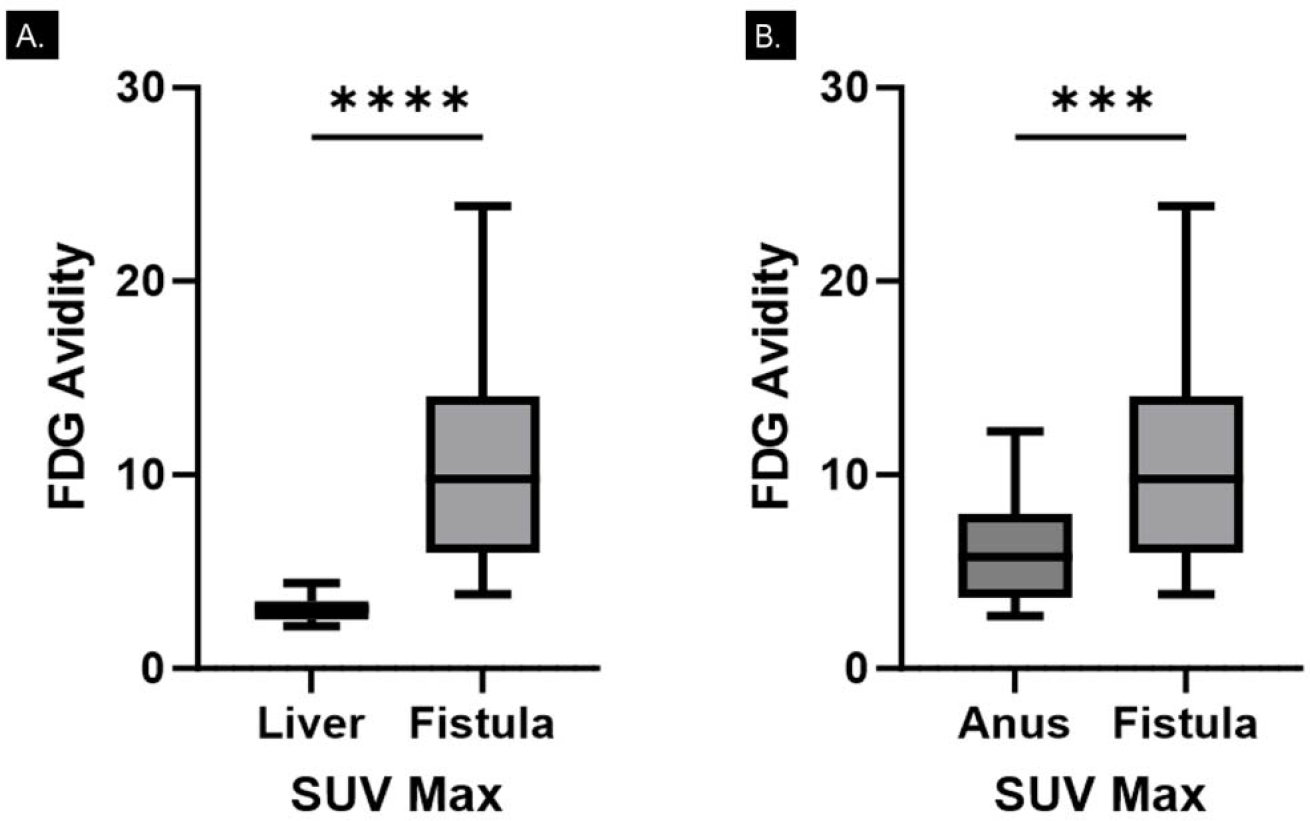
Difference in FDG avidity between fistula and liver in A (p<0.0001) and fistula and anus (p=0.0005) in B.

**Figure 3.**
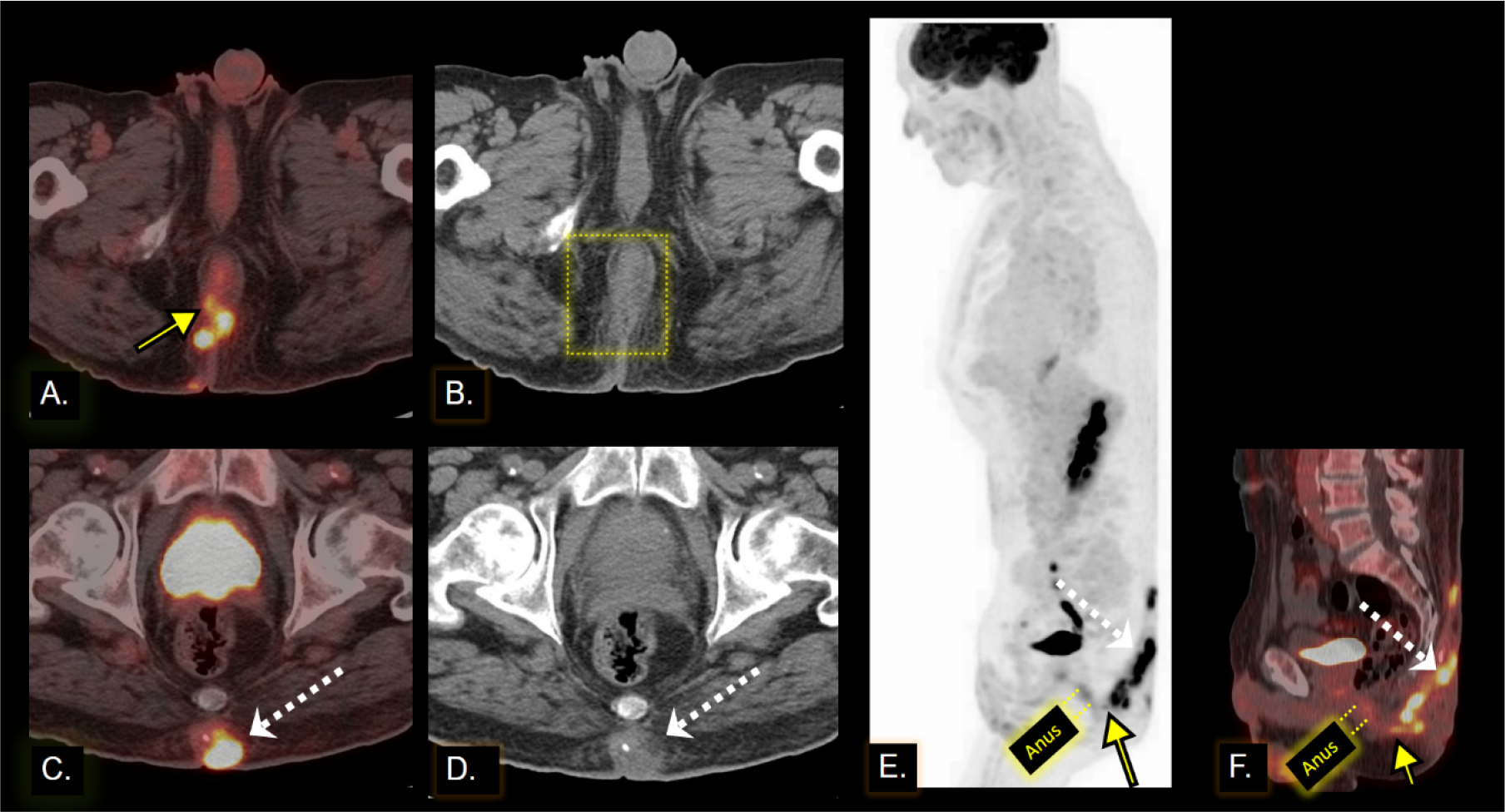
Patient undergoing staging for esophageal cancer with an incidental finding of increased FDG activity in a perianal fistula portrayed through (A, C) axial and (F) sagittal projections of fused PET/CT images along with (B, D) non-fused axial CT images and (E) a sagittal maximum intensity projection. (A) The perianal fistula (yellow arrow) is apparent on the fused FDG PET/CT and demonstrates an FDG avid tract protruding from the anal verge. (B) On review of the non-contrast CT images, there is no identifiable correlate or essential abnormality other than some loss of subcutaneous fat and diffuse stranding in the right gluteal cleft fat surrounding the anal verge (yellow dotted box). The fistula feeds an abscess (white dotted arrows in C, D, E, and F) centered in the gluteal cleft fat, which tracks cranially, resulting in abscess pooling around the area of the subcutaneous back fat overlying the coccyx, as demonstrated by the linear ascending FDG avidity in E and F. On the non-contrast axial CT image in D, the hyper-attenuating rounded soft tissue collection met our study criteria for an abscess, as it measured greater than 1 centimeter in maximum trans-axial diameter, up to 1.9 centimeter. This is best classified as a Saint James grade 2 fistula, which is a perianal fistula feeding a branching tract and resulting in a pooling abscess in the subcutaneous fat. Sagittal reconstructions of the maximum intensity projection and fused FDG-PET/CT in E and F demonstrate the relationship of the anal canal uptake (between the yellow dotted lines) to the fistula (yellow arrow) exiting at the anal verge.

SUV_max_ for all regions of interest along with SUV_max_ ratios of the fistula and the anus are recorded in **Table 2**. The average SUV_max_ ratios of fistula-to-liver were 3.41 (range: 1.76-6.14) and 3.54 (range: 1.11-6.42) for the anorectal cancer group and non-anorectal cancer group, respectively, without a significant difference between the two groups (p=0.86). There were no significant differences in other FDG-avidity ratios between the anorectal and non-anorectal groups. Interestingly, mean (±SD) anal SUV_max_ for the non- anorectal cancer group (5.69±2.42) was not significantly different from the anorectal cancer group (6.67±3.16) (p=0.42) (**Table 3**).

**Table 2.** Summary statistics for all patients.

| | Total, n | Mean ± SD |
| --- | --- | --- |
| **Fistula SUV<sub>max</sub>** | 24 | 10.8 ± 5.28 |
| **Abscess SUV<sub>max</sub>** | 6 | 15.8 ± 4.91 |
| **Anus SUV<sub>max</sub>** | 24 | 5.98 ± 2.63 |
| **Rectum SUV<sub>max</sub>** | 24 | 5.14 ± 3.7 |
| **Liver SUV<sub>max</sub>** | 24 | 3.09 ± 0.584 |
| **Fistula-to-anus SUV<sub>max</sub> ratio** | 24 | 2.12 ± 1.39 |
| **Anus-to-liver SUV<sub>max</sub> ratio** | 24 | 1.85 ± 0.608 |
| **Fistula-to-liver SUV<sub>max</sub> ratio** | 24 | 3.5 ± 1.6 |

**Table 3.** Comparing mean SUV_max_ values between anorectal and non-anorectal groups.

|  | **Anorectal cancer group, n** | **Anorectal cancer group Mean ± SD** | **Non-anorectal cancer group, n** | **Non-anorectal cancer group Mean ± SD** | **p value** |
| --- | --- | --- | --- | --- | --- |
| **Fistula SUV<sub>max</sub>** | 7 | 9.03 ± 3.45 | 17 | 11.5 ± 5.81 | 0.31 |
| **Abscess SUV<sub>max</sub>** | 2 | 15.4 ± 6.9 | 4 | 15.9 ± 4.92 | 0.92 |
| **Anus SUV<sub>max</sub>** | 7 | 6.67 ± 3.16 | 17 | 5.69 ± 2.42 | 0.42 |
| **Rectum SUV<sub>max</sub>** | 7 | 7.07 ± 5.18 | 17 | 4.34 ± 2.7 | 0.1053* |
| **Liver SUV<sub>max</sub>** | 7 | 2.8 ± 0.414 | 17 | 3.2 ± 0.613 | 0.13 |
| **Fistula-to-anus SUV<sub>max</sub> ratio** | 7 | 1.56 ± 0.883 | 17 | 2.34 ± 1.52 | 0.23* |
| **Anal-to-liver SUV<sub>max</sub> ratio** | 7 | 2.02 ± 0.518 | 17 | 1.78 ± 0.641 | 0.38 |
| **Fistula-to-liver SUV<sub>max</sub> ratio** | 7 | 3.41 ± 1.85 | 17 | 3.54 ± 1.54 | 0.86 |

\*Non-normal distribution.

8 patients had a St. James grade 1 fistula. The mean fistula SUV_max_ for St. James grade 1 fistulas was 8.04±6.61. 6 patients had a St. James grade 2 fistula. The mean fistula SUV_max_ for St. James grade 2 fistulas was 12.6±3.57. 3 patients had a St. James grade 3 fistula. The mean fistula SUV_max_ for St. James grade 3 fistulas was 9.32±5.63. 4 patients had a St. James grade 4 fistula. The mean fistula SUV_max_ for St. James grade 4 fistulas was 12.1±3.82. 3 patients had a St. James grade 5 fistula. The mean fistula SUV_max_ for St. James grade 5 fistulas was 10.9±3.72 (**Table 4**). Fistula-to-liver SUV_max_ ratios were significantly lower for patients with St. James grade 1 fistulas (2.34±1.35) than those with grade 2 (4.24±1.32; p=0.0224) and grade 4 (4.38±1.61; p=0.0295) fistulas, but was not significantly different from grade 3 (2.84±0.829) and grade 5 (3.8±1.93) fistulas (**Figure 4**). All other comparisons were not significantly different.

**Table 4.**
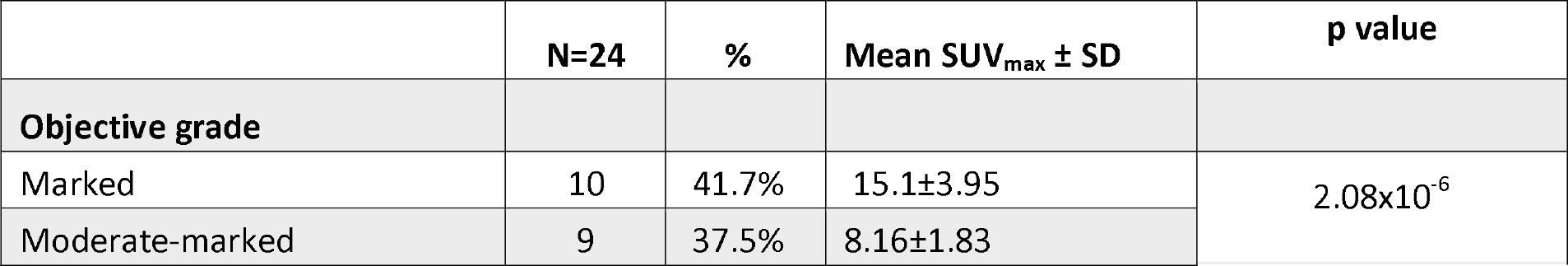
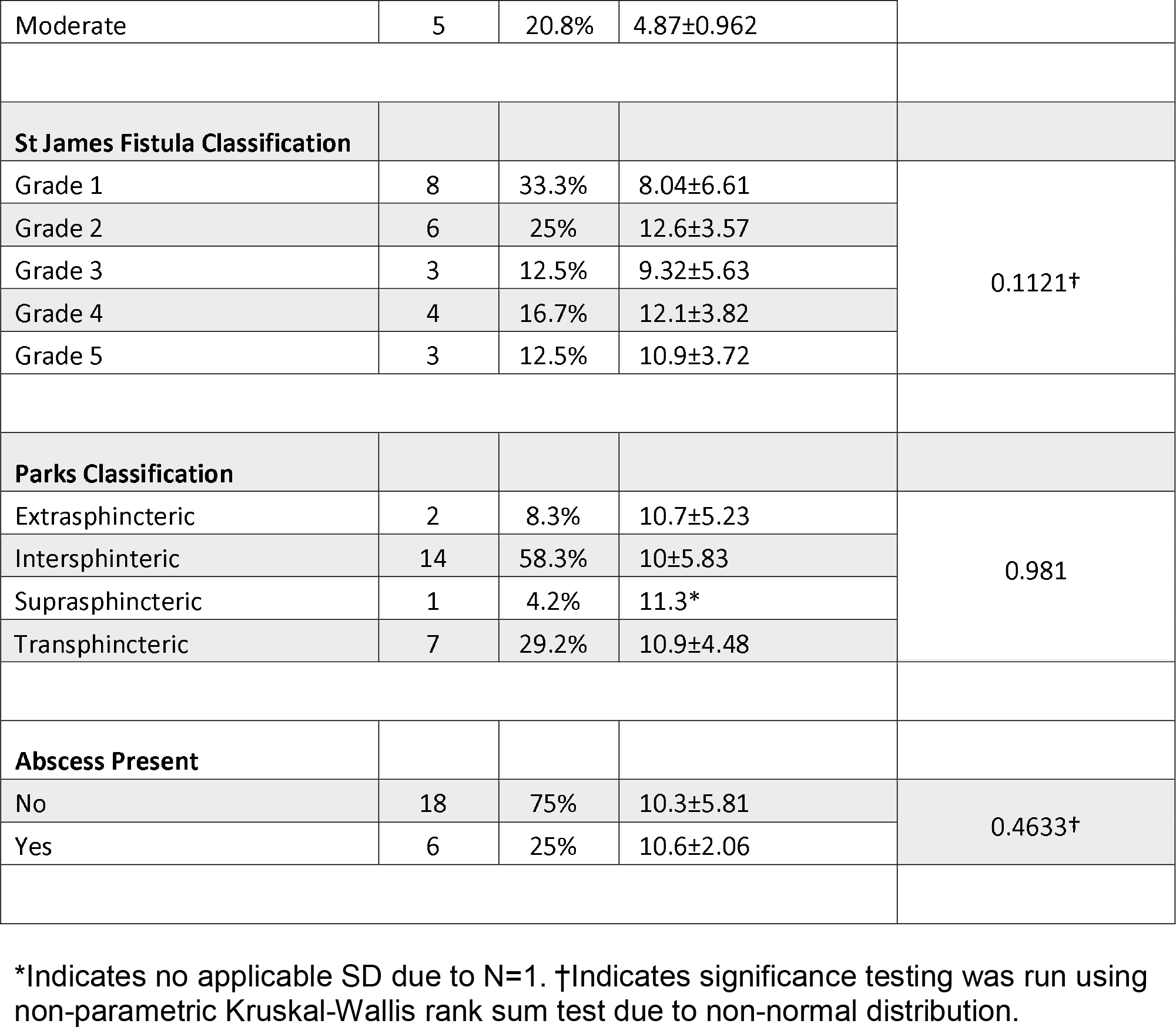
Mean fistula SUV_max_ values based on objective grading, St. James grading, Parks Classification, and presence of abscess.

|  | **N=24** | **%** | **Mean SUV<sub>max</sub> ± SD** | **p value** |
| --- | --- | --- | --- | --- |
| **Objective grade** |  |  |  |  |
| Marked | 10 | 41.7% | 15.1±3.95 | 2.08x10<sup>-6</sup> |
| Moderate-marked | 9 | 37.5% | 8.16±1.83 |  |
| Moderate | 5 | 20.8% | 4.87±0.962 |  |
| **St James Fistula Classification** |  |  |  |  |
| Grade 1 | 8 | 33.3% | 8.04±6.61 | 0.1121† |
| Grade 2 | 6 | 25% | 12.6±3.57 |  |
| Grade 3 | 3 | 12.5% | 9.32±5.63 |  |
| Grade 4 | 4 | 16.7% | 12.1±3.82 |  |
| Grade 5 | 3 | 12.5% | 10.9±3.72 |  |
| **Parks Classification** |  |  |  |  |
| Extrasphincteric | 2 | 8.3% | 10.7±5.23 | 0.981 |
| Intersphincteric | 14 | 58.3% | 10±5.83 |  |
| Suprasphincteric | 1 | 4.2% | 11.3* |  |
| Transphincteric | 7 | 29.2% | 10.9±4.48 |  |
| **Abscess Present** |  |  |  |  |
| No | 18 | 75% | 10.3±5.81 | 0.4633† |
| Yes | 6 | 25% | 10.6±2.06 |  |

\*Indicates no applicable SD due to N=1. †Indicates significance testing was run using non-parametric Kruskal-Wallis rank sum test due to non-normal distribution.

**Figure 4.**
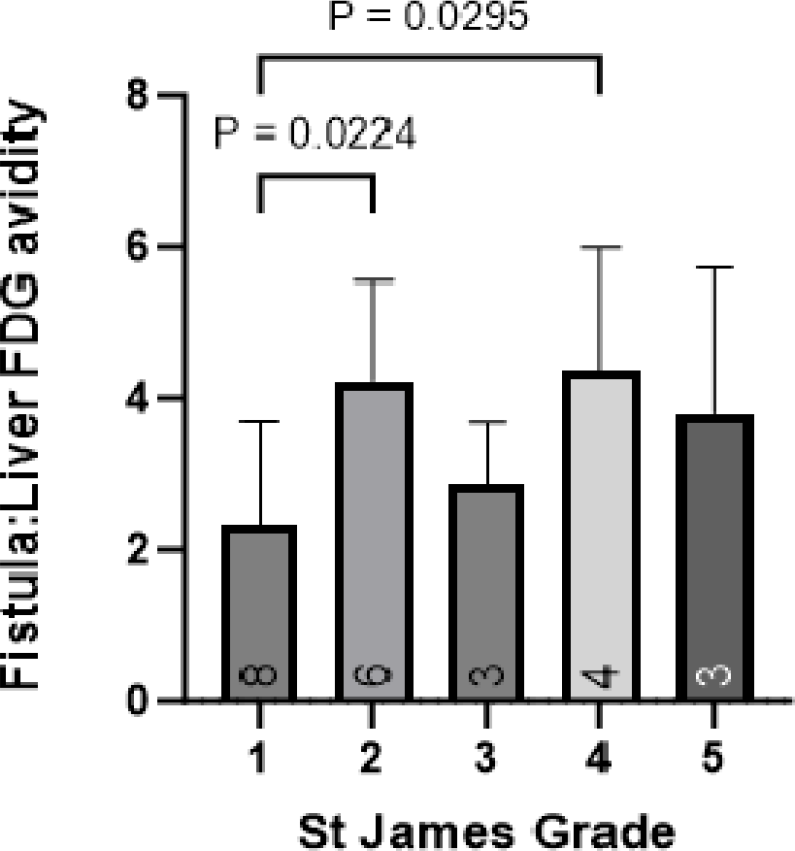
Comparing FDG activity between different types of fistulas based on St. James grading scale. Significant differences between St. James grade 1 vs. all other grades and St. James grade 1 vs. St. James grade 2 are denoted by p value brackets. All other comparisons were not statistically significant.

Although the mean SUV_max_ ratio for St. James grade 3 was lower than grades 2-5, this was not significantly different. Examples of each grade of fistula on FDG-PET/CT images are shown in **Figure 5**. The number of patients with extrasphincteric, intersphincteric, suprasphincteric, and transphincteric fistulas per Parks classification was 2, 14, 1, and 7 respectively. The mean fistula SUV_max_ for extrasphincteric fistulas was 10.7±5.23. The mean fistula SUV_max_ for intersphincteric fistulas was 10±5.83. The fistula SUV_max_ of the singular suprasphincteric fistula was 11.3. The mean fistula SUV_max_ for transphincteric fistulas was 10.9±4.48. There was no significant difference in SUV_max_ ratios between intersphincteric and transphincteric fistulas per Parks classification. Mean fistula SUV_max_ values based on objective grading, St. James grading, Parks classification, and presence of abscess are depicted in **Table 4**.

**Figure 5.**
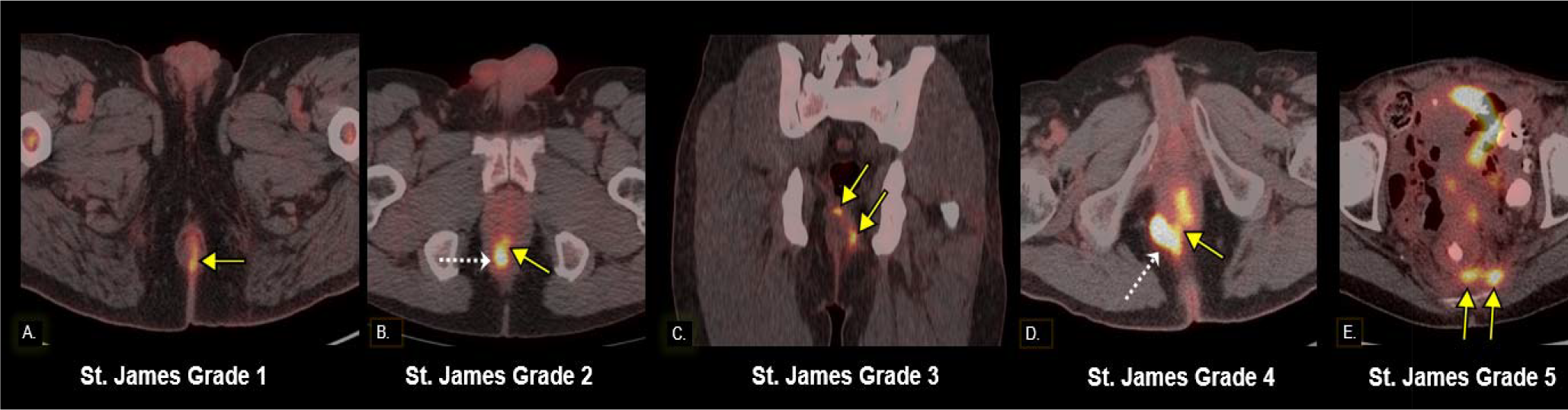
Examples of fused FDG-PET/CT manifestations of the St. James grading system for perianal fistulas. (A) Axial image of mildly FDG avid fistulous tract (yellow arrow) protruding out of the anal verge. (B) Axial image of an FDG avid tract (yellow arrow) which protrudes from the anal verge and feeds a small abscess (dashed white arrow). (C) Coronal image of a linear FDG avid tract (yellow arrows) which extends through the internal sphincter and protrudes externally, creating a simple trans- sphincteric pathway. (D) Axial image of an FDG avid fistula tract (yellow arrow) which violates the external sphincter and feeds an FDG avid right ischiorectal abscess (dashed white arrow). (E) Axial image of a suprasphincteric fistula (yellow arrows) that feeds into the left mesorectal fascia and eventually protrudes through the left levator ani.

## Discussion

In this descriptive study consisting of patients undergoing FDG-PET/CT for oncologic diagnosis and staging, we identified those with incidental perianal fistulas or abscesses and aimed to quantify fistula FDG uptake in relation to the anal canal and liver. We found that FDG uptake within perianal fistulas is significantly higher than FDG uptake in the liver and anal canal, with an average fistula SUV_max_ value of 10.8. This increased uptake reflects the heightened metabolic activity in perianal fistulas due to inflammation [14]. This suggests that PET can be used to identify fistulas as discrete from surrounding structures such as the anal canal. Assessing the level of inflammation on PET via visualization and SUV_max_ quantification can provide additional functional information that could enhance the identification of fistulas and abscesses, which may appear equivocal on a non-contrast CT alone, similar to the case demonstrated in **Figure 3**. This underscores the potential of PET/CT as a diagnostic tool for detecting fistulas in patients with contraindications to MRI and contrast-enhanced CT. Although FDG-PET/CT may not be the primary diagnostic modality for perianal fistulas, recognizing them as incidental findings can equip radiologists and nuclear medicine physicians with the confidence to identify these lesions during PET interpretation.

Our subgroup analysis comparing SUV_max_ values and ratios between patients with and without anorectal cancer demonstrated no significant difference between both groups. This may suggest that the level of inflammation within perianal fistulas in patients with anorectal cancer is comparable to that of patients without anorectal cancer. It may also be due to limited ability to discover differences between both groups due to low power of the study. There was some variation in FDG uptake within fistulas based on St.

James grading, with St. James grade 1 fistulas demonstrating lower uptake compared to higher-grade fistulas, which postulates that higher-grade fistulas may be associated with higher level of inflammation and metabolic activity, or that higher grade fistulas have larger diameters given SUV_max_ dependency on lesion size. The lack of significance when comparing St. James grades 2-5 fistulas is likely due to the small number of patients in each group. Regardless of significance, it is understandable that mean SUV_max_ ratio for St. James grade 3 is lower than grade 2 and grade 4 fistulas because both grade 2 and 4 fistulas are associated with an abscess which have higher lesion sizes and levels of inflammation that could contribute to higher SUV_max_ values. Although the differences between all groups were not statistically significant, further research can be undertaken to assess the ability of PET/CT to assist with providing fistula classification information prior to further management.

To the best of our literature search, no published studies have investigated FDG uptake in perianal fistulous tracts. One study investigated incidental non-malignant causes of FDG uptake, and a perianal fistula with marked uptake accounted for an incidental finding in 1 of 212 patients [15]. In another study, patients with incidental anal FDG uptake on PET/CT underwent a follow-up anorectal exam and a fistula was recorded as one of the diagnoses [16]. Patients with perianal fistulizing Crohn’s disease are at risk for cancer development in long-standing fistulous tracts, and few studies have noted FDG uptake in fistulous tracts with malignancy [17, 18].

Some studies have assessed FDG uptake in normal anal canals and other non-malignant anal processes like hemorrhoids. In one study which involved the retrospective analysis of PET/CT scans to determine FDG uptake in the anal canal of 201 healthy patients, 15.4% of patients demonstrated high FDG uptake in the anal canal during the early phase of imaging. The average anal SUV_max_ in the early phase was 3.82 for these patients, significantly lower than the average for anal cancer patients [19]. Another study compared the SUV_max_ of the anal canal in patients with and without hemorrhoids and discovered a mean SUV_max_ of 2.9 (range: 1.4-8.3) in patients with hemorrhoids [8]. To the best of our literature search, our study is the first to quantify anal FDG uptake in patients with fistulas. The average anal SUV_max_ in the non-anorectal cancer group was 5.69, which was slightly lower when compared to the anorectal cancer group with an average of 6.67. However, the difference was not significant. These findings indicate that increased FDG uptake in the anal canal does not necessarily suggest malignancy.

This study has several inherent limitations. Limited sample size may impact the statistical power and generalizability of the results. Due to the retrospective study design, patient data were obtained at varying stages of hematologic/oncologic treatment and the quality of clinical reference standards was not uniform across the patient cohort which limited our ability to control for potential confounding factors. In this study, our focus was on physician confirmatory diagnosis of perianal fistula or abscess, and dedicated follow-up and fistula-related outcomes were not systematically assessed. Serial follow-up of fistulas or concordance with other imaging modalities was not systematically evaluated. Although future research should address these important follow-up factors, we were not able to assess them in this study due to the heterogeneity of available data and variations in the timing of follow-up. Instead, this study’s main objective was to provide radiologists with insights into the one-time diagnosis of perianal fistula or abscess, aiming to assist in interpreting this incidental finding on PET/CT scans. Perianal abscesses also demonstrated markedly increased FDG avidity with an average abscess SUV_max_ of 15.8. Due to the small number of abscesses identified in this study, we were unable to perform meaningful statistical analysis for comparison between patients with and without abscesses.

## Conclusion

This study provides evidence of increased FDG activity within perianal fistulas, indicative of heightened metabolic activity in these regions due to inflammation. FDG uptake in the fistula was significantly higher than that of the liver and anal canal without a significant difference between patients with and without anorectal cancer. Furthermore, our results propose that an elevated FDG uptake in the anal canal does not necessarily indicate the presence of anal cancer, a significant consideration for radiologists and clinicians. Overall, our findings highlight the potential of PET/CT as a diagnostic tool in detecting perianal fistulas and abscesses in patients with contraindications to MRI or contrast-enhanced CT, highlighting the need for continued research in this direction.

## Data Availability

All data produced in the present study are available upon reasonable request to the authors.

## References

1. Garcia-Olmo, D., et al., Prevalence of anal fistulas in Europe: systematic literature reviews and population-based database analysis. Advances in therapy, 2019. 36(12): p. 3503–3518.

2. Singh, K., et al., Magnetic resonance imaging (MRI) evaluation of perianal fistulae with surgical correlation. Journal of clinical and diagnostic research: JCDR, 2014. 8(6): p. RC01.

3. Levy, A.D., et al., ACR Appropriateness Criteria® Anorectal Disease. J Am Coll Radiol, 2021. 18(11s): p. S268–s282.

4. Guniganti, P., et al., Imaging of acute anorectal conditions with CT and MRI. Abdom Radiol (NY), 2017. 42(2): p. 403–422.

5. Caliste, X., et al., Sensitivity of computed tomography in detection of perirectal abscess. Am Surg, 2011. 77(2): p. 166–8.

6. Meisner, R.S., et al., Pilot study using PET/CT as a novel, noninvasive assessment of disease activity in inflammatory bowel disease. Inflamm Bowel Dis, 2007. 13(8): p. 993–1000.

7. Holtmann, M.H., et al., 18F-Fluorodeoxyglucose positron-emission tomography (PET) can be used to assess inflammation non-invasively in Crohn’s disease. Dig Dis Sci, 2012. 57(10): p. 2658–68.

8. Tsai, S.C., et al., Findings of 2-fluoro-2-deoxy-d-glucose positron emission tomography in hemorrhoids. Abdom Imaging, 2011. 36(5): p. 548–51.

9. Wahl, R.L., V. Dilsizian, and C.J. Palestro, At Last, (18)F-FDG for Inflammation and Infection! J Nucl Med, 2021. 62(8): p. 1048–1049.

10. Jamar, F., et al., EANM/SNMMI guideline for 18F-FDG use in inflammation and infection. J Nucl Med, 2013. 54(4): p. 647–58.

11. Morris, J., J.A. Spencer, and N.S. Ambrose, MR imaging classification of perianal fistulas and its implications for patient management. Radiographics, 2000. 20(3): p. 623–35; discussion 635-7.

12. Parks, A.G., P.H. Gordon, and J.D. Hardcastle, A classification of fistula-in-ano. Br J Surg, 1976. 63(1): p. 1–12.

13. Holleran, S. and R. Ramakrishnan, cufunctions, a package to facilitate statistical analyses in R. 2021, New York, NY: Columbia University Medical Center.

14. Włodarczyk, M., et al., Current concepts in the pathogenesis of cryptoglandular perianal fistula. J Int Med Res, 2021. 49(2): p. 300060520986669.

15. Metser, U., et al., Benign nonphysiologic lesions with increased 18F-FDG uptake on PET/CT: characterization and incidence. AJR Am J Roentgenol, 2007. 189(5): p. 1203–10.

16. Moussaddaq, A.-S., et al., Incidental anal 18fluorodeoxyglucose uptake: Should we further examine the patient? World Journal of Clinical Cases, 2020. 8(17): p. 3679.

17. Ogawa, H., et al., Adenocarcinoma associated with perianal fistulas in Crohn’s disease. Anticancer Res, 2013. 33(2): p. 685–9.

18. Sasaki, H., et al., Clinicopathological characteristics of cancer associated with Crohn’s disease. Surg Today, 2017. 47(1): p. 35–41.

19. Sena, Y., et al., Physiological 18F-FDG uptake in the normal adult anal canal: evaluation by PET/CT. Ann Nucl Med, 2020. 34(8): p. 538–544.

